# User-Centered Design to Develop and Implement an ML-Based Asthma Management Tool

**DOI:** 10.1101/2022.12.28.22282986

**Authors:** Lu Zheng, Joshua W. Ohde, Shauna M. Overgaard, Tracey A. Brereton, Kristelle A. Jose, Chung-Il Wi, Kevin J. Peterson, Young J. Juhn

## Abstract

**Background:** Personalized asthma management depends on a clinician’s ability to efficiently review patient’s data and make timely clinical decisions. Unfortunately, efficient and effective review of these data is impeded by the varied format, location, and workflow of data acquisition, storage, and processing in the electronic health record. While machine learning and clinical decision support tools are well-positioned as potential solutions, the translation of such frameworks requires that barriers to implementation be addressed in the formative research stages. Transparency, accountability, suitability, and adaptability may be bolstered by clinician engagement through a direct empathetic approach aimed at determining complex user requirements of implementation, usability, and workflow integration.

**Objectives:** We aimed to utilize a structured user-centered design approach (double-diamond design framework) to 1) qualitatively explore clinicians’ experience with the current asthma management system, 2) identify user requirements to improve algorithm explainability and A-GPS prototype, and 3) identify potential barriers to ML-based CDS system use.

**Methods:** At the ‘discovery’ phase, we first shadowed to understand the practice context. Then, semi-structured interviews were conducted online with 14 clinicians who provide asthma care at two outpatient facilities. Participants were asked about their current difficulties in gathering information for pediatric asthma patients, their expectations of ideal workflows and tools, and suggestions on user-centered interfaces and features. At the ‘define’ phase, a synthesis analysis was conducted to converge key results from interviewees’ insights into themes, eventually forming critical ‘how might we’ research questions to guide model development and implementation.

**Results:** We identified user requirements and potential barriers associated with three overarching themes: 1) Usability and Workflow Aspects of the ML System, 2) User Expectations and Algorithm Explainability, and 3) Barriers to Implementation in Context. Even though the responsibilities and workflows vary among different roles, the core asthma-related information and functions they requested were highly cohesive, which allows for a shared information view of the tool. Clinicians hope to perceive the usability of the model with the ability to note patients’ high risks and take proactive actions to manage asthma efficiently and effectively. For optimal machine-learning algorithm explainability, requirements included documentation to support the validity of algorithm development and output logic, and a request for increased transparency to build trust and validate how the algorithm arrived at the decision. Acceptability, adoption, and sustainability of the asthma management tool are implementation outcomes that are reliant on the proper design and training as suggested by participants.

**Conclusions:** As part of our comprehensive informatics-based process centered on clinical usability, we approach the problem using a theoretical framework grounded in user experience research leveraging semi-structured interviews. Our focus on meeting the needs of the practice with machine learning technology is emphasized by a user-centered approach to clinician engagement through upstream technology design.

## Introduction

Transparency, suitability, and adaptability are cited reasons for the chasm between advances in artificial intelligence (AI) and implementation in health systems ^1^. Hindering implementation is a lack of transparency about the data used to make decisions and recommendations ^2^. The conceptual suitability of, or aversion to, an algorithm in clinical use is practically governed by a clinician’s autonomous decision to engage with the tool ^3^. The adaptability of the algorithm to local patient populations and unique workflows further increases the likelihood of adoption ^4^. It is logical that a proactive and systematic approach to addressing barriers to transparency, suitability, and adaptability may propel the wider implementation and adoption of AI in patient care ^5^.

Ultimately, the foundation of this approach is rooted in clinician engagement at the earliest stages of AI development ^6^. Determining the user’s complex and diverse requirements for effective Machine Learning-based Clinical Decision Support (ML-based CDS) tools requires a thorough understanding of the clinical utility of data sources and suitable designs to facilitate contact and response in appropriate settings ^7^. This formative usability approach can be achieved through an empathetic and sustained relationship within a multidisciplinary team initiated by early-stage formative research and upstream technology design ^8^.

In a personalized medical practice aiming to optimize a clinician’s management of asthma, efficient review of the condition’s characterizing features is critical ^9^. Unfortunately, efficient and effective review of these data using electronic health records (EHR) and timely clinical decisions are impeded by the varied format, location, and workflow of data acquisition, storage, and processing ^10^. To support clinicians, our aim is to develop an ML-based CDS tool that 1) predicts future risk of asthma exacerbation (risk stratification and resource management), 2) provides this risk evaluation in the context of a summary of relevant information for asthma management (reduction of EHR review burden), and 3) offers options for actionable intervention.

Described in our published work, our AI evaluation plan employs a phased framework (Figure 1) to address technical performance, usability and workflow, and health impact, and iteratively follows our model documentation steps ^11^. In this paper, we address Phase I: Technical performance and safety, highlighting user experience (UX) design research through stakeholder shadowing and interviews.

**Figure 1.**
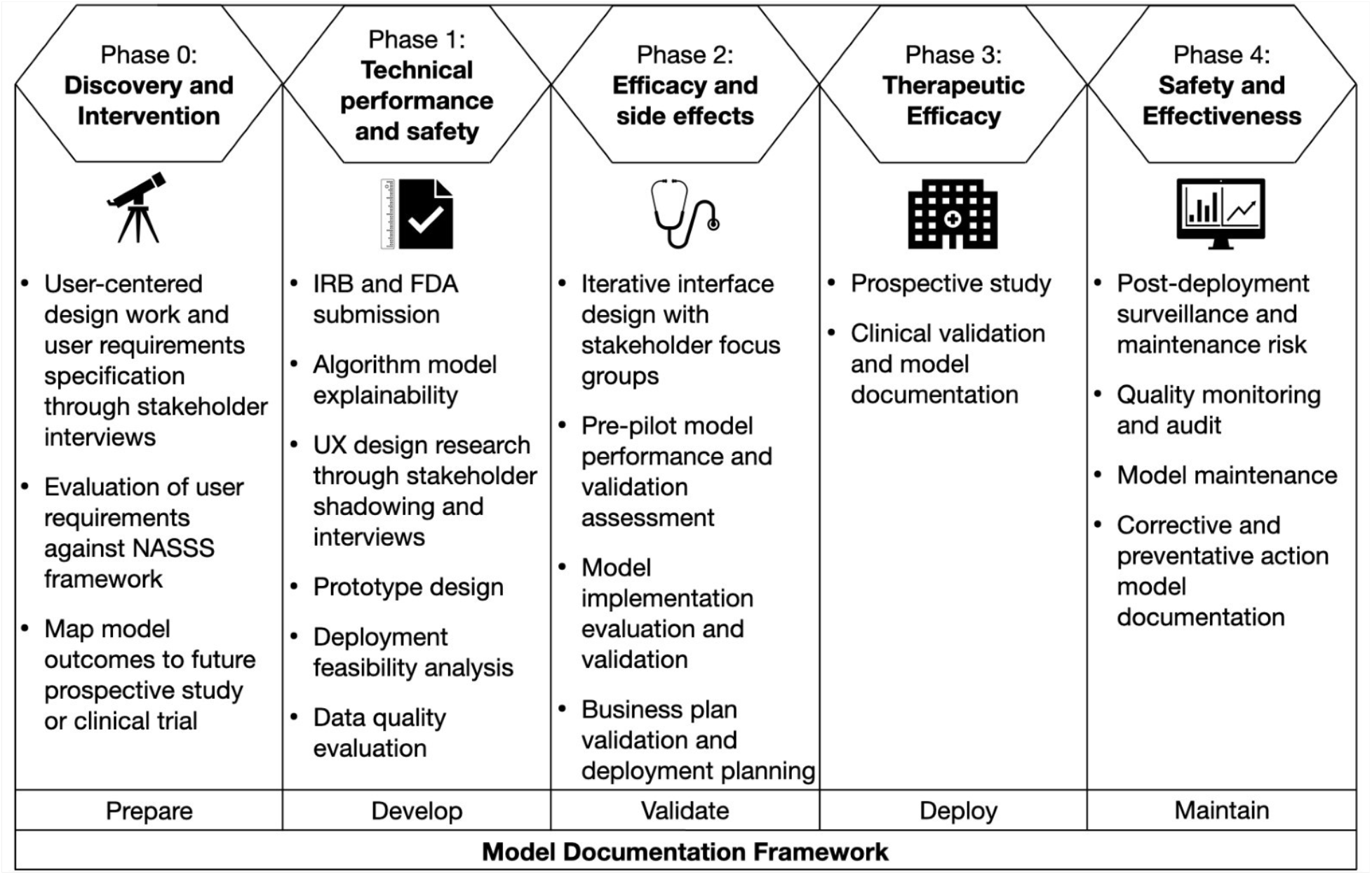
Phased research framework for evaluation of AI applied to the Asthma Guidance and Prediction System (A-GPS) project based on Park et al. (2020), with the internally developed model documentation framework ^12^.

### Theoretical Framework

Literature detailing ML-based CDS tool translation indicates an ineffective balance of tool intelligence with explainability, creating gaps in translation, implementation, and accountability ^13^. This suggests the need for early engagement with clinical stakeholders to mitigate present gaps by gaining a comprehensive and multidisciplinary understanding of present challenges, as well as identifying requirements for development and integration that prioritize both intelligence and practical usability. UX research methodologies were adopted throughout Phase 1: Technical performance and safety, including the Double Diamond design framework and participatory design method that strategically engaged clinical stakeholders to derive unmet needs and identify user requirements ^14,15^. The Double Diamond design framework (Figure 2), a graphical guide following the phases of the design process, was applied to customize and standardize progression of UX research by incorporating iterative loops and feedback opportunities to progress development ^16,17^. This framework supports human-centered design, specifically participatory design, an approach that invites stakeholders to be active in the design process as a means of better understanding, meeting, and preempting needs to inform developer efforts ^6^. At the ‘research’ stage, the UX research team conducted interviews and gathered data detailing challenges and pain points of current pediatric asthma care processes, as well as ideas for innovation. At the ‘synthesis’ stage, the UX research team categorized data gathered from interviews into themes and reframed findings into opportunities in the form of How-Might-We (HMW) questions ^18^. Derived HMW questions served as actionable prompts that acknowledge current challenges requiring solutions and encourage collaborative solution generation representative of relevant clinician’s experiences^18,19^. This UX design process allowed for the communication of user requirements from the perspective of engaged stakeholders to provide direct guidance and inform tool development, thereby moving past present challenges of trying to design for stakeholders and beginning to design with them at the ‘ideation’ phase ^20^.

**Figure 2.**
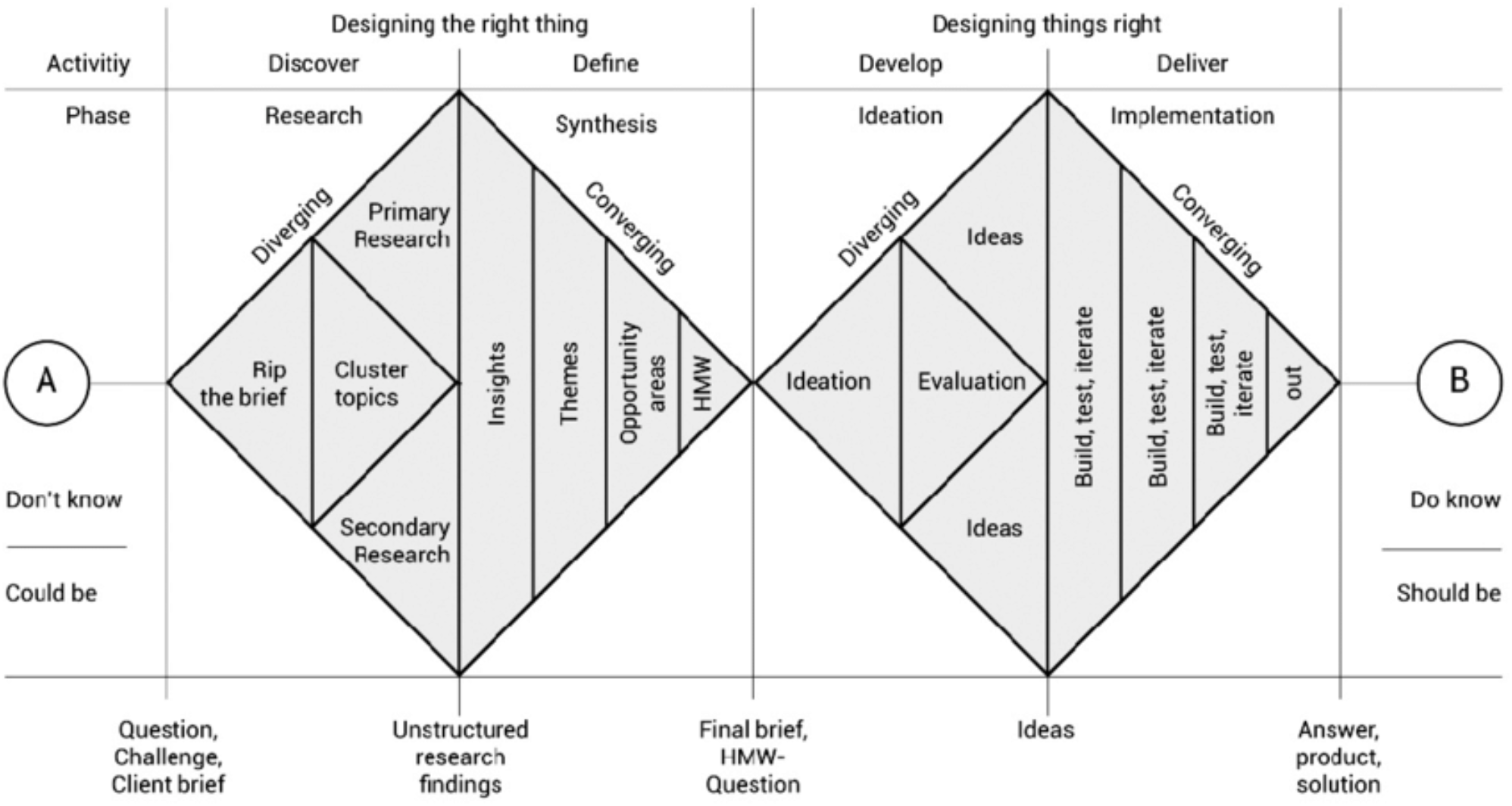
The Double Diamond design framework^21^, supplemented with a participatory design method, provided a framework for UX research to follow in Phase 1, promoting active engagement of stakeholders in tool development.

### Study Objectives

The Asthma Guidance and Prediction System (A-GPS) tool is an ML-based CDS tool accessible from the EHR. It aims to summarize all asthma-related context information extracted from the EHR on one screen page^9,22^. The tool will be embedded with a functional component of the Asthma Exacerbation risk model (AE risk model), which applies ML algorithms to predict patient’s risk of exacerbation in one year ^23^. The study objectives were to 1) qualitatively explore clinicians’ experience with the current asthma management system, 2) identify user requirements to improve algorithm explainability and A-GPS prototype, and 3) identify potential barriers to ML-based CDS system use. Research questions were developed to probe challenges and pain points of gathering asthma-related information within the current asthma management system, thorough evaluation of clinical team member workflows, user requirements for prototype optimization, algorithm explainability and display.

## Methods

### Participant Selection and Recruitment

We invited a group of clinicians representing the key roles in asthma management (primary care practice leadership, primary care physicians, primary nurse practitioners, asthma care coordinators, and asthma specialty practice lead for primary care) from the Department of Pediatric and Adolescents Medicine (DPAM) and Family Medicine outpatient practices to help assess various aspects of implementation of a new tool in practice. Ideally, the potential users of A-GPS, such as pediatricians, care coordinators, and project stakeholders could be included. A total of 14 participants were recruited by email using a convenience sampling approach and scheduled for a one-on-one, 30 to 60-minute virtual interview.

### Data Collection

This formative usability research was directed toward an understanding of user requirements and to facilitate optimal workflow integration, estimate the potential impact of healthcare delivery factors, and work capacity constraints on achieved benefit. We aimed to collect different facets of qualitative data to identify all stakeholders, understand user needs, probe for optimal tool design to support clinical decision-making and routine workflow for each group in a comprehensive manner. To get the clinical context of how the tool will be used in practice, one researcher shadowed both sites and described the general patient flow. Next, we scheduled a 60-minute virtual interview with each recruited participant. An introductory statement provided background on the ML-based CDS tool prototype and explained the goal of the interview, developing rapport with interview participants. Each interview session was composed of two parts. Part one was a routine 30-minute semi-structured interview. Interview guides were created for stakeholders, clinicians that were part of the A-GPS project or practice leadership, such as a division or practice chair, and users, defined as those with no stake in A-GPS but are practicing clinicians. Detailed interview guides were attached as supplementary material (Appendix 1). Stakeholders were asked specific questions regarding their role as stakeholders in A-GPS and as potential users. Non-stakeholder participants (users) were questioned about their experience and needs as end users. Within part two of an interview session, participants were invited to demonstrate an EHR walkthrough on their working computer. During this time, we observed how the clinician routinely uses the system and defines the asthma-related information required to make a medical decision. Additionally, follow-up questions were asked to explore their cognitive process. While part one was focusing on clinicians’ reported problems and individual opinions, part two allowed us to observe the current problems and workflow objectively. Each type of data supplemented the other to achieve problem-probing and user needs consolidation.

### Data Analysis

User experience (UX) specialized translational informaticians engaged with practice components to evaluate usability and workflow to determine effectiveness, efficiency, satisfaction, ease of use, explainability, and utilization, as described in the AI Evaluation Framework by Overgaard et al. 2022^23^. Interviews were transcribed, reviewed, and coded by team members LZ, JO, KJ, and TB. Using an online collaboration tool MURAL (Tactivos, Inc.)^24^, transcripts were coded by identifying emergent themes and categorized into primary research questions asking what, how, where, and when. Other themes included challenges/pain points, barriers to adoption, novel ideas, new insights, and stakeholder considerations. Sub-themes were presented as opportunities for change using a HMW question format ^18^. Figure 3 provides a brief look into the synthesis and analyzing work completed utilizing the MURAL tool. As for the EHR walkthrough, we used the data as a reference to make the list of the acquired information in the EHR system.

**Figure 3.**
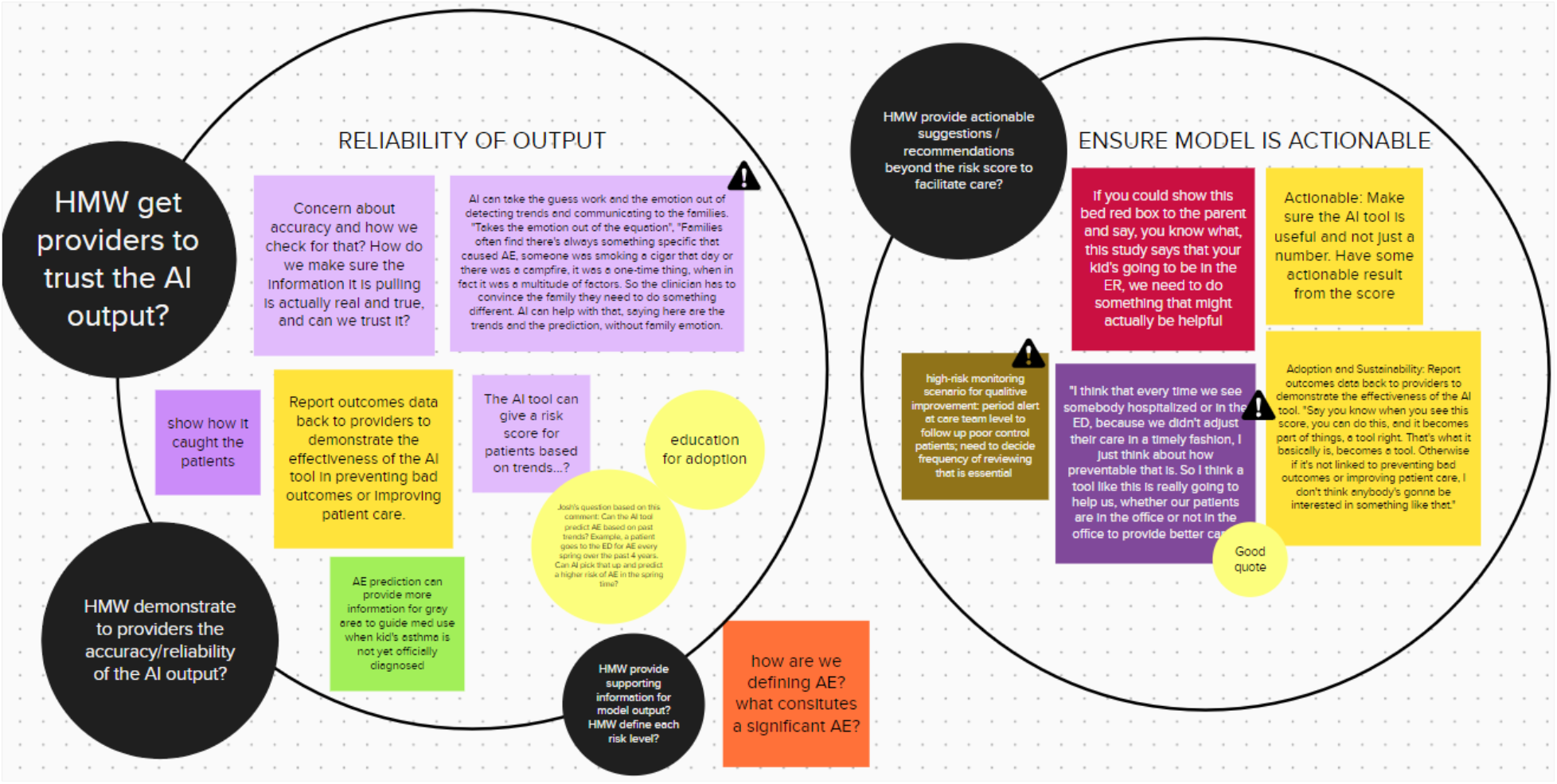
MURAL (Tactivos, Inc.) is an online collaborative tool. Key quotes from participant transcripts were added to the board, utilizing colors to identify participants. Similar or consistent responses across participants were grouped together within circles, as shown in the image, to form sub-themes.

## Results

### Participant Characteristics

A total of 14 clinicians were interviewed across four Mayo Clinic Health System sites in Minnesota, including Rochester, Red Wing, Albert Lea/Austin, and Kasson. Of those who participated, 7 (50%) were female, 11 (79%) were physicians (MD or APRN) and 3 (21%) were nurses. Their roles included asthma specialist, allergist, pulmonologist, pediatrician, family medicine physician, asthma care coordinator, and resident.

### Targeted Patient Population and Clinician Users of A-GPS Tool

Clinicians identified opportunities to enhance asthma management for the diagnosed and at-risk population through use of the A-GPS tool. According to clinicians, the tool would best serve pediatric patients with a diagnosis of asthma and should also aim to cover pediatric patients not officially diagnosed but at high risk of developing asthma, including those identified with the following conditions: symptoms of wheezing or coughing, albuterol or oral steroids use, frequently reported respiratory conditions of pneumonia, lung infections, wheezing, or coughing.

> *There are patients who probably have asthma that we don’t detect, but that’s where I think this tool would be helpful because maybe even though they don’t have a diagnosis of asthma, they’ve had wheezing, or other things listed in their diagnosis and problem list. That would be helpful to avoid missing those people. [P6]*
>
> *Some kids had been given a bronchodilator because often at 18 months, they present with like viral induced wheezes, and we find it improves with albuterol. So, we get a response to albuterol, and we know that these kids are potentially likely to get asthma, but we typically don’t make that diagnosis until after two. [P4]*

Pediatric asthma patients may be seen by multidisciplinary clinical roles including allergy specialists, pulmonologists, pediatricians, asthma care coordinators, rooming nurses, ED physicians, and primary care providers. When identifying proper clinician users of the tool, it was reported that any role that needs to provide asthma management care in practice would benefit from accessing and utilizing A-GPS tool. To capture potential role-based variance in user requirements, the routine workflows and information needs were asked for each participant. It was found that asthma management is coordinated care by dynamic care teams, however, participants demonstrated preference for a shared view of the tool to gain a shared understanding of patient cases. Even though the responsibilities and workflows vary among different roles, the core asthma-related information and functions they requested were highly cohesive, which allows for a shared information view of the tool.

### Usability and Workflow Aspects of ML System

In general, clinicians welcomed the integration of the AE risk model into the patient’s EHR. In practice, the prediction results are expected to help facilitate preventive actions to support better asthma management. To accomplish this, the AE risk prediction results cannot simply be in the EHR, it also needs to notify clinicians and prompt the care team to follow up with patients in an expedited fashion.

> *If risk prediction results are added and approaching the threshold, and you will get out a message letting you know that that is happening, that would be the best way to go. Because then you can prevent the next exacerbation, rather than waiting to see the patient next time they come to clinic, by then patient might have been through a couple of exacerbations. [P2]*
>
> *If we are getting this risk score and especially if it were telling them that this is somebody that is at high risk of relapses and recurrences of episodes, then we can make that effort to reach out to those individuals. That should be flowing in my mind. That should be going to our care teams. [P14]*

Despite the goal of being proactive, getting clinician’s attention to the right patient at the right time in an acceptable format is an issue. Notification methods were suggested by participants and opinions varied based on roles.

> *When you open the chart, it be helpful to have that notification sent via an in-basket message so that we’re aware and could follow up sooner. There might be cases where we’re aware that they’re high risk, but we can just delete it if we already have that plan for follow-up. [P9]*
>
> *Best Practice Advisory (BPA) kind of prompts the provider that some action needs to be taken in these areas. And it might be a nice opportunity. Or one of the things we have is emergency action plans. Some similar way that incorporates into an action that needs to be taken or addressed for this patient. [P6]*

In-basket message was mentioned by many clinicians as a common type of active alarm. But it is necessary to balance effective information delivery and alert fatigue as clinicians, especially physicians, receive various alarms and notifications from multiple channels in their daily work.

> *I would just like the color coding in the records. I do not know if an in-basket message would be effective because we get a lot of them. If it was, I like the message was really clear and can quickly know what it is for. [P13]*
>
> *Probably not an alarm for high-risk cases. I can imagine people getting annoyed at that, but if it came up in care gaps on the story board, like a reminder of something needing to be reviewed, that would be nice. [P4]*
>
> *I hate to say in-basket messages because that just generates another inbox that the provider doesn’t have time to handle. I think having these folks show up as high-risk followed up by our care coordinators is the right way of handling this. It should be a trigger to get care coordinators or nurses to schedule a visit with the patient, which is more important than notifying the provider. [P14]*

Additionally, the alarm or notification should reach clinicians with proper guidance for next steps.

> *In BPA, we see alarms as this bright red thing with exclamation points. We’re going to want to act on that, but how do you act on it? Like, does it prompt then if you go into your plan, will it prompt something where you get some choices, like high-risk, you know. Whatever the risk score is, here’s some options for you and you click those, and it goes into an order set, and you can order it and you’re done. [P7]*

### User expectations and algorithm explainability

#### Expectations and Perceived Impact

Participants reported that the A-GPS tool is expected to have a positive impact on clinicians’ workflow and patient experiences. Clinicians anticipate a positive impact on usability and workflow by 1) streamlining the review of asthma information, 2) providing patients with “proactive” rather than “reactive” care, 3) empowering patients with a deeper and more personalized understanding of their condition, and 4) improving outcomes. Participants find the tool can be helpful in several situations, such as preparing for an upcoming patient visit, following up on a patient’s conditions remotely, and changing or refilling medications based on changes in a patient’s condition. With well-organized asthma information presented at the appropriate time in the workflow, clinicians expect they can save time reviewing information and the care team will have a consistent understanding of patient cases.

Importantly, proactive and preventive care is anticipated with the AE risk model, allowing the prioritization of resources to patients of greatest need, and reduction of AE, ED visits, and hospitalizations. Clinicians hope to perceive good usability of the model with the ability to note patients’ high risks and take proactive actions to manage asthma efficiently and effectively. Ideally, with attention-grabbing model output visualization, both patient and caregiver would be more engaged in home-based care after seeing future risks. The potential to further drive higher quality outcomes was identified in the potential to monitor the relationship between patient adherence to medication, symptoms, and other contributing factors.

#### Algorithm Explainability

User requirements of the asthma exacerbation prediction algorithm output emphasized interpretability, logical justification, and validation. Specifically, known definitions and levels of risk categories must be explainable, leading to efficient patient classification and resource allocation. Visual indications of severity, such as red, yellow, and green to define high, medium, and low-risk categories paired with a numerical indication were required. Supporting contextual information such as flagging primary features impacting risk prediction and providing a summary of additional asthma management variables were key requirements. Supporting information should be easily accessible and presented as hovering capabilities or links to relevant data (e.g., patient history, baseline diagnostics). To assist with algorithm explainability and informing next steps, users required supplemental information on how the prediction score was calculated, bolstered by comparative diagnostics (e.g., individual and population baseline values). Clinicians expressed concern regarding accuracy and reliability without significant validation of the model. Requirements included documentation to support the validity of algorithm development and output logic, and a request for increased transparency to build trust and validate how the algorithm arrived at the decision. For successful integration, users require that strategic education and phased implementation must be offered. Education and regular reports on the clear demonstration of value was the preferred strategy to gain an understanding of appropriate A-GPS use and limitations. Examples of stated learning preferences included hands-on training, such as workshops presented at monthly meetings, regular follow-up communication and showcasing of successful use cases, and video tutorials. Importantly, users require a clear demonstration of value to ease adoption, achieved by a phased implementation approach (multi-site).

> *I want to be able to see that risk score. When the patient is in front of me, I also want to be able to see a whole lot more information about that patient, preferably in an easy to find format that I don’t have to go digging in Epic for it, like I currently do. [P14]*
>
> *I would probably like something simpler, like not necessarily a percentage. And then I like, okay, it’s red, which means they’re at a high risk. In the background, I could know what that means. And if you want more information, then you could click and find why it is high. [P13]*
>
> *I think high, medium, low would, you know, would be sufficient. And if you would have something popped up or even color coded too, like they are low risk in green, medium in yellow. If they’re high risk and in red, that certainly will get your attention. I also want to know what is putting them at risk. Is it severity of symptoms, their need for oral steroids, their hospitalization and ED visits? So that would certainly be helpful to know exactly where their risk area is. [P9]*
>
> *I would like it to give me a percentage score rather than a level. So, if I had a model, I would like to see a percentage within a certain period of time. Like within two years or within one year, there is a percentage chance that there’ll be an asthma exacerbation. Some people like the simplicity of a one to five. But if I’m making my decisions, I kind of want a little bit more detail on it. And also, if I click on it, I would like to be able to see where and what data points they’re using to make these predictions because sometimes the data in the Epic chart is incorrect. [P4]*

### Barriers to Implementation in Context

#### Accuracy and Reliability

Many participants expect the AI model given by A-GPS will be validated for accuracy and reliability. They also stressed the importance of making the model explainable and transparent to users. Clearly explaining why the model predicted a specific risk score will allow users to understand the logic of the model and its relevance to the patient’s current asthma situation. Without demonstrating validation or providing transparency clinicians will lack trust in the tool and likely not use it, limiting its clinical value.

#### Ethical Considerations

Clinicians recognized the potential benefits of A-GPS but voiced several ethical concerns regarding the AE risk model. One concern was the misunderstanding of AI’s role in clinical practice and that AI will override clinician autonomy to make clinical decisions. However, one participant asserted that the goal of AI is to provide complementary information and that the clinician would still make the final clinical decisions. A similar concern was the impact the risk prediction model would have on a clinician’s intuition. More specifically, when the AE risk model contradicts the clinician’s professional judgment, the possibility of legal or ethical issues may arise depending on what action the clinician takes.

> *Machine learning introduces a new wrench in things. Because now you’re not giving me a necessarily a recommendation, but you’re giving me insight that might either raise my intuition or lower it. How do you handle having that prediction result legally and ethically and everything else? [P12]*
>
> *As a pulmonologist, I am trying to understand how other systematic diseases impact their asthma. So, I am also checking tests of other body systems and evaluating by talking to patients. For the populations I am seeing with asthma, hopefully at some point artificial intelligence could help us, but I just do not see it at this point. [P2]*
>
> *This prediction score is not meant to override. This is complimentary information for you. I know you do mental calculations, but this is a data driven calculation that gives you other complimentary information. If there’s a discrepancy, is there anything you are thinking low in emotion, say ‘hi, just to think about it on this page’. So then, you know, you don’t have to go to that page, just look through another page of sectional summary. [P5]*

A patient-specific concern was the potential for unnecessary anxiety and emotional burden on patients and their caregivers when told the AE risk model deems the child at “high risk”. The fear that an asthma event could occur based on a prediction tool that many patients and caregivers may not fully understand may provoke unnecessary changes in the child’s daily activities, as shown in the example below.

> *Parents may worry about their child if the AI tool said, “high risk of AE” and subsequently change daily decisions, such as not sending their child to school or letting them play outside. [P7]*

## Discussion

Principal results are discussed by identified themes. In each theme, we started with “How Might We” questions to inspire discussions on challenges and opportunities.

### Usability and Workflow Aspects of the ML System

Challenges and opportunities of usability and workflow aspects of the AI system prompt key questions such as: *How might we incorporate the A-GPS tool to support workflows of different roles?* Clinicians are tasked with a workload that involves increasing patient volumes, more complex diseases, and an overwhelming EHR system. One goal of A-GPS is to help alleviate the time clinicians spend in the EHR to find asthma-related information and supplement the clinical decision process required to minimize a patient’s risk of AE. Successfully incorporating A-GPS into the current workflow of various clinical roles is arguably as important as the tool itself. Participants suggested placing the A-GPS tool in the same location within the EHR and having the same view, regardless of roles. This will allow easier navigation in the EHR within the care team’s current practice as clinicians, nurses, and care coordinators frequently view each other’s screens during patient care.

### User expectations and algorithm explainability

Challenges and opportunities of user expectations and algorithm explainability prompt key questions such as: *1) How might we communicate A-GPS results in a way that is explainable to patients?* Although patients and caregivers were not interviewed, they will receive some level of information from the A-GPS tool communicated to them by the clinician or care team. The way in which the outcome of the AE risk model is explained to patients will be important. Limiting unnecessary anxiety or misunderstandings while still effectively communicating the model results needs to be carefully addressed so families can make appropriate decisions that improve the patient’s outcomes. Properly educating clinicians on how to explain the AE risk model to families is one approach that could be tied into the overall education plan for A-GPS. *2) How might we remove barriers to adoption to increase clinician buy-in?* The adoption of any new tool or technology rarely goes as planned, it takes time to achieve buy-in from users. To increase clinicians’ buy-in for A-GPS a few barriers should be addressed. First, clinicians need to see that the A-GPS tool is validated, accurate, reliable, and effective at saving time in the EHR. Communicating this data using various educational modalities can increase the reach among clinicians. Second, some clinicians will wait to see the value A-GPS brings to their colleagues before using it themselves. These individuals may be reached by leveraging clinician champions who believe the A-GPS tool improves user experience with the EHR and patient outcomes. Ultimately, the best method to facilitate adoption of the A-GPS tool is to ensure its functionality meets user’s needs and expectations. Clinicians are more likely to use A-GPS if a clear and concise ML-based CDS tool is created that contains only asthma-related information with easy access to more detailed notes and test results. Moreover, clinicians may use and act on the AE risk model if they trust it, understand what the risk score means for their patient, and understand how the model came to its conclusion for their patient.

### Barriers to Implementation in Context

Challenges and opportunities related to implementation and system use in context prompt key questions such as: *How might we address ethical issues brought on by a difference between the AE risk model and a clinician’s professional judgment?* The contradiction between the AE risk model and a clinician’s clinical assessment may pose an ethical and even legal issue. In practice, clinicians may feel pressured to act upon the model’s output in fear of legal challenges even if they believe the patient’s risk of AE is different based on their professional judgment. Although this topic deserves further exploration, it is reasonable to assume that educating clinicians that their clinical judgment supersedes the result from the AE risk model as the model does not take into consideration the multitude of variables the clinician assesses. Moreover, the reason for the development of the AE risk model in this context was to provide supplemental information to improve the care of patients with asthma, not replace the expertise of clinicians. Addressing this upfront with potential users should be a component of A-GPS implementation.

Acceptability, adoption, and sustainability of A-GPS are implementation outcomes that are reliant on the proper design and training as suggested by participants. Without following the guidance of clinicians given in this study the success of A-GPS will be limited, resulting in decreased user satisfaction and clinical effectiveness. To overcome potential barriers to implementation success there are several priority areas that should be met. First, the A-GPS tool needs to be easily accessible within the EHR, in a location that is obvious and consistent across all clinical roles, contains all asthma-related information on a single page, and is visually concise and intuitive. To increase acceptability, the AE risk model needs to be validated for accuracy and reliability and made transparent to users. Transparency is necessary to build trust among clinicians and trust facilitates acceptance and adoption. The AE risk model output needs to be easily interpretable, clearly defined, and intuitive to improve adoption and sustainability. Except for the quality of model output, the importance of quality and strategy of education and training cannot be ignored. In a recent paper by Gordon et al., Mayo Clinic took a standardized and efficient approach to provide education and training sessions when implementing a new EHR system^25^. The results demonstrated higher acceptance and confidence among users. This could be a good example for A-GPS project in terms of successful implementation.

### Strengths and Limitations

Conducting interviews with potential users of a new clinical tool not only gains insights into their needs, but also encourages buy-in as seeking their input prior to implementation demonstrates that the research team values user feedback. To our knowledge, this study facilitated buy-in and support among participants as several thanked the study team for their efforts to understand user needs. The translational informatics and data science teams engaged clinical stakeholders in creating materials to ensure appropriate interpretation, contextual information, and educational support for use of the model. UX research was leveraged to engage with clinical stakeholders and prospective end-users by gaining a comprehensive and multidisciplinary understanding of the role A-GPS is expected to play in pediatric asthma care. Shadowing and interviewing clinical stakeholders were a source of engagement that gathered user requirements from the perspective of potential users, with the objective of informing tool development and translation efforts ^20^. Once A-GPS tool is functioning technically, its fit into the clinical workflow must be evaluated. Moreover, education and documentation must be provided to explain the algorithm and its limitations to effectively translate between the perspectives of experts that created and support the technology and the perspectives of experts that employ the solution to patients. Evaluating the interpretation needs of clinicians, preferences for the display of model output (e.g., percentage vs. binary threshold), and feature contributions will be assessed based on the data obtained from UX research efforts. Concurrently, the team will also engage clinician stakeholders in the development of model documentation to support explainability ^26^. The data obtained from UX research will assist the translational informatics and data science teams in identifying the level of explainability needed to inform and validate the design of A-GPS and supplementally enhance existing workflows ^27^. Strategic efforts to promote explainability include applying a documentation framework grounded in scientific research addressing known challenges. This encompasses interdisciplinary best practice reporting requirements that follow phases of model development (Prepare, Develop, Validate, Deploy, Maintain) for knowledge continuity throughout the solution’s life cycle ^11^.

A limitation of this study is its generalizability as participants were selected using a convenience sampling approach and may not be representative of all potential A-GPS users. Though, we included stakeholders from Mayo Clinic Rochester, academic practice with teaching hospital and Mayo Clinic Health System to represent a real-world practice in community and maximize the generalizability of findings. Another limitation is that the perspective of patients and their caregivers were not evaluated in this study. This was purposeful as the intended users of A-GPS are clinicians but the impact the AE risk model may have on families, as stated in the Ethical Considerations section, should be explored further.

### Further Research

Future research will evaluate the sustainability and scalability of user requirements for enterprise, national, and international adoption of the ML-based CDS tool. Ethical considerations of AI interpretation, patient engagement, and clinician autonomy warrant further investigation. Our research team will conduct multiple studies as we approach the future stages of Efficacy and Side Effects, Therapeutic Efficacy, and Safety and Effectiveness planned in our phased comprehensive AI evaluation framework (Stage I). There are more questions to answer in the future: *How might we demonstrate to providers the accuracy and reliability of the AI output? How might we define required transparency for AI output? How might we provide an efficient educational module for users and show validation measures to support and explain AI output? How might we handle alarm fatigue, including situations where patients do not respond to providers’ intervention? How might we improve ML-model predicting the risk of AE when the provider’s proper and timely intervention may reduce the performance of the model (e*.*g*., *positive predictive value)?*

### Conclusions

We aimed to anticipate barriers to the translation of our pediatric asthma management ML-based CDS tool by engaging clinicians in prototype development and optimization leveraging UX research methodologies. In efforts to bolster the transparency, suitability, and adaptability of our solution we qualitatively evaluated user requirements and potential barriers in three overarching themes: usability and workflow aspects of the ML system, user expectations and algorithm explainability, and barriers to implementation in Context. We presented findings specific to our tool’s risk evaluation in the context of a summary of relevant information for asthma management. This work contributes to Phase I of our comprehensive informatics-based AI evaluation framework developed by our multidisciplinary team of clinicians, data scientists, translational informaticians, and UX experts at Mayo Clinic ^23^. The transparent evaluation and documentation of AI applications in healthcare enhances clinician and patient trust, supports sharing of AI between hospitals, and increases standards and shared responsibility across the continuum of care. The results of this development study further enhance the model documentation of A-GPS aimed to ensure rigorous evaluation, transparency, and knowledge continuity ^11^. A sustainability and scalability evaluation of user requirements will strengthen the potential for national and international adoption of A-GPS.

## Supporting information

Supplemental Interview Guide

## Data Availability

All data produced in the present study are available upon reasonable request to the authors

## Acknowledgements

We would like to deeply thank Dr. Manuel Arteta, Dr. Valeria Cristiani, Joy Fladager Muth, Dr. Gregory Garrison, Dr. Margret Gill, Dr. Jason Greenwood, Darcy Hall, Dr. Martha Hartz, Dr. Julian Lynaugh, Dr. Donnchadh O’Sullivan, Dr. Sarah Scherger, Dr. Eric Schnaith and Dr. David Soma for finding their time for interview and sharing their valuable insights into usability of A-GPS in clinical practice setting. This project was supported by the Center for Digital Health, Precision Population Science Lab, the AI Program of Department of Pediatric.

