## Supplemental Interview Guide for "User-Centered Design to Develop and Implement an ML-Based Asthma Management Tool"

**Appendix 1. Interview Guides**

Stakeholder Questions

1. Tell us about your role in the project.
2. What pain points are you trying to solve in the project?
3. What gap are you trying to fill?
4. How was the project idea formed initially?
5. Why do you think developing such a tool is a workable solution in asthma care?
6. Who are the clinical users of the tool in practice?
7. Which patient population can benefit from the project?
8. What actions do you expect users to take when a patient is identified as “high risk” by the tool?
9. If we implement the tools in your department, what are the potential logistical or conceptual challenges we need to overcome?
10. What do you think about sustainability and future opportunities of the tool?
11. What does long term success of the project look like to you?

Current Problem Questions

1. Tell us about your role in a patient’s asthma management.
2. Describe a situation where you typically review a patient’s asthma-related information.
3. How do you pull asthma-related information from EHR?
4. What information is the most important for asthma-related treatment plans?
5. What decision do you need to make when reviewing the information?
6. What are the challenges with finding the information in the current system?
7. How long do you estimate it takes to collect all the needed information for asthma management from the current system?
8. What do you think can be improved?

A-GPS Tool Ideation Questions

1. What are your initial thoughts/impressions of A-GPS?
2. What are your intuitive expectations?
3. Describe your ideal workflow experience with the tool.
4. When and where do you think the tool should be implemented? Why?
5. In your role, what are the top 5 most important items you would like to see?
6. How would you like to see this information presented?
7. How can we provide sufficient support (training, education resources, help tools) providers?
8. What are the barriers do you anticipate in the adoption of this tool?

Asthma Exacerbation Risk Model Questions

1. What would you expect from the ML model tool?
2. When and where do you want to see the risk model results?
3. How would you like to see the model output displayed?
4. How would you interpret the clinical meaning of ML model outputs?
5. What information do you think would help you better understand the risk results to achieve explainability, transparency, and accountability?
6. How will the ML output change what you do clinically for a patient?
7. What actions will you take for high-risk patients?
8. What kind of support is necessary for successful adoption?
9. What do you think are the barriers affecting the model results being utilized?
